# The first 6 weeks – setting up a UK urgent dental care centre during the COVID-19 pandemic

**DOI:** 10.1101/2020.05.06.20093179

**Authors:** Emily Carter, Charlotte C Currie, Abisola Asuni, Rachel Goldsmith, Grace Toon, Catherine Horridge, Sarah Simpson, Christopher Donnell, Mark Greenwood, Graham Walton, Ben Cole, Justin Durham, Richard Holliday

**Affiliations:** Newcastle Hospitals NHS Foundation Trust, Newcastle upon Tyne, UK; School of Dental Sciences, Newcastle University, Newcastle upon Tyne, UK

**Author notes:** Joint first author (Emily Carter, Charlotte C Currie).

## Abstract

**Introduction:** The COVID-19 pandemic has posed many challenges, including provision of urgent dental care. This paper presents a prospective service evaluation during establishment of urgent dental care in the North-East of England over a six-week period.

**Aim:** To monitor patient volumes, demographics and outcomes at the North-East urgent dental care service and confirm appropriate care pathways.

**Main Outcome Methods:** Data were collected on key characteristics of patients accessing urgent care from 23^rd^ March to 3rd May 2020. Analysis was with descriptive statistics.

**Results:** There were 1746 patient triages, (1595 telephone and 151 face-to-face) resulting in 1322 clinical consultations. The most common diagnoses were: symptomatic irreversible pulpitis or apical periodontitis. 65% of clinical consultations resulted in extractions, 0.5% an aerosol generating procedure. Patients travelled 25km on average to access care, however this reduced as more urgent care centres were established. The majority of patients were asymptomatic of COVID-19 and to our knowledge no staff acquired infection due to occupational exposure.

**Conclusion:** The urgent dental care centre effectively managed urgent and emergency dental care, with appropriate patient pathways established over the 6-week period. Dental preparedness for future pandemic crisis could be improved and informed by this data.

**Three In Brief Points:**

1. A summary is given of how urgent dental care was established in the North East of England during the COVID-19 pandemic which may help with future preparedness for pandemics.
2. Aerosol generating procedures were almost always avoided in the delivery of urgent dental care
3. A telephone triage system was effectively used to determine who needed clinical care, and to separate symptomatic, asymptomatic and shielding patients, with very few failures in triage noted.

## Introduction

The COVID-19 pandemic is likely to be one of the biggest challenges to face medicine and dentistry globally over the last century. During the week of 16^th^ March 2020, the impact of COVID-19 began to affect dentistry in the UK. Primary dental care services were advised to radically reduce and subsequently stop routine dental care, patient face-to-face contact, and arrangements had to be made for the provision of care in urgent dental care clinics (UDCCs) during the pandemic.

The Newcastle Dental Hospital (NDH) is part of the Newcastle upon Tyne Hospitals (NUTH) NHS Foundation Trust and serves a population of over 3 million (1). It delivers specialist dental services alongside undergraduate and postgraduate training and is fully integrated with the School of Dental Sciences at Newcastle University. The Dental Emergency Clinic and Child Dental Health Department at NDH have been providing a walk-in clinic for many years with the primary purpose of enabling clinical teaching for undergraduate dental students and training postgraduate dentists, whilst providing a valuable service for the local population.

During the pandemic the dental emergency clinic was adapted to provide multidisciplinary seven-day emergency and urgent dental care (2) to all age groups and was staffed by qualified dentists from within the hospital, including Consultants, specialists, specialty trainees and dental core trainees in Oral Surgery, Restorative Dentistry, Paediatric Dentistry, Special Care/Community Dentistry and Orthodontics with support from Oral and Maxillofacial Surgery Consultants. At this early stage and in the absence of national guidance, the clinic had to balance providing a core service for the most in need (aiming to minimise dental attendances at Medical Emergency Departments) against limiting patient throughput with a new, stricter triaging system to protect staff, other patients, and to preserve potentially limited personal protective equipment (PPE) supplies. Following the inception of the adapted clinic on the 23^rd^ March 2020, national guidance (3) was published on the 25^th^ March, stipulating that dental practices stop seeing patients face-to-face, leading to 337 NHS practices in our region closing their doors overnight. This guidance stated for patients to contact their General Dental Practitioner (GDP) for Advice, Analgesia and/or Antimicrobials (commonly referred to as AAA) in the first instance, prior to any referral to a local UDCC to try and minimise patient travel and unnecessary attendances.

The urgent dental care (UDC) service at the NDH evolved rapidly in response to challenges and changes in service and policy, whilst remaining open to the public. Figure 1 illustrates some key events in the COVID-19 pandemic alongside milestones in the development of the NDH UDCC. The clinic quickly transitioned to using remote triage for all patients, both in, and out-of-hours (OOH). In-hours (Monday-Friday 8.30-16.30) triage was initially via walk-in and direct calls to the clinic from patients and dentists. With the assistance of NHS111, a national free-to-call non-emergency medical helpline (4), the in-hours service at NDH progressed to only accepting referrals by email, mainly from GDPs and NHS111 to manage the flow of referrals. A limited number of exceptions were made for clinically urgent walk-in cases who were triaged at a designated desk at the entrance of the building. All patients referred by email had a further telephone triage by a dentist to establish their need for treatment and advice or an appointment was given. OOH triage (i.e. weekends) was provided by NHS111/DCAS. Both in-hours and OOH the service was divided into three distinct areas within the hospital: one for COVID-19 symptomatic and isolating patients; another for the asymptomatic; and a separate clinic for shielding and vulnerable patients (referred to as shielding in our practice) to minimise the risk of cross infection. The criteria for COVID-19 symptomatic patients was in line with NHS advice (5) and we limited treatment in this group to true dental emergencies only (trauma, swelling or bleeding). The criteria for shielding patients is defined by the NHS as those who would be ‘extremely vulnerable’ if they were to contract COVID-19. These patients were grouped with those defined as ‘vulnerable’ under the general headings of: immunocompromised, severe respiratory disease, type I diabetes, pregnant, over the age of 70 and ‘other’. Appropriate PPE was used for all staff in line with local and national guidance (6–9).

**Figure 1.**
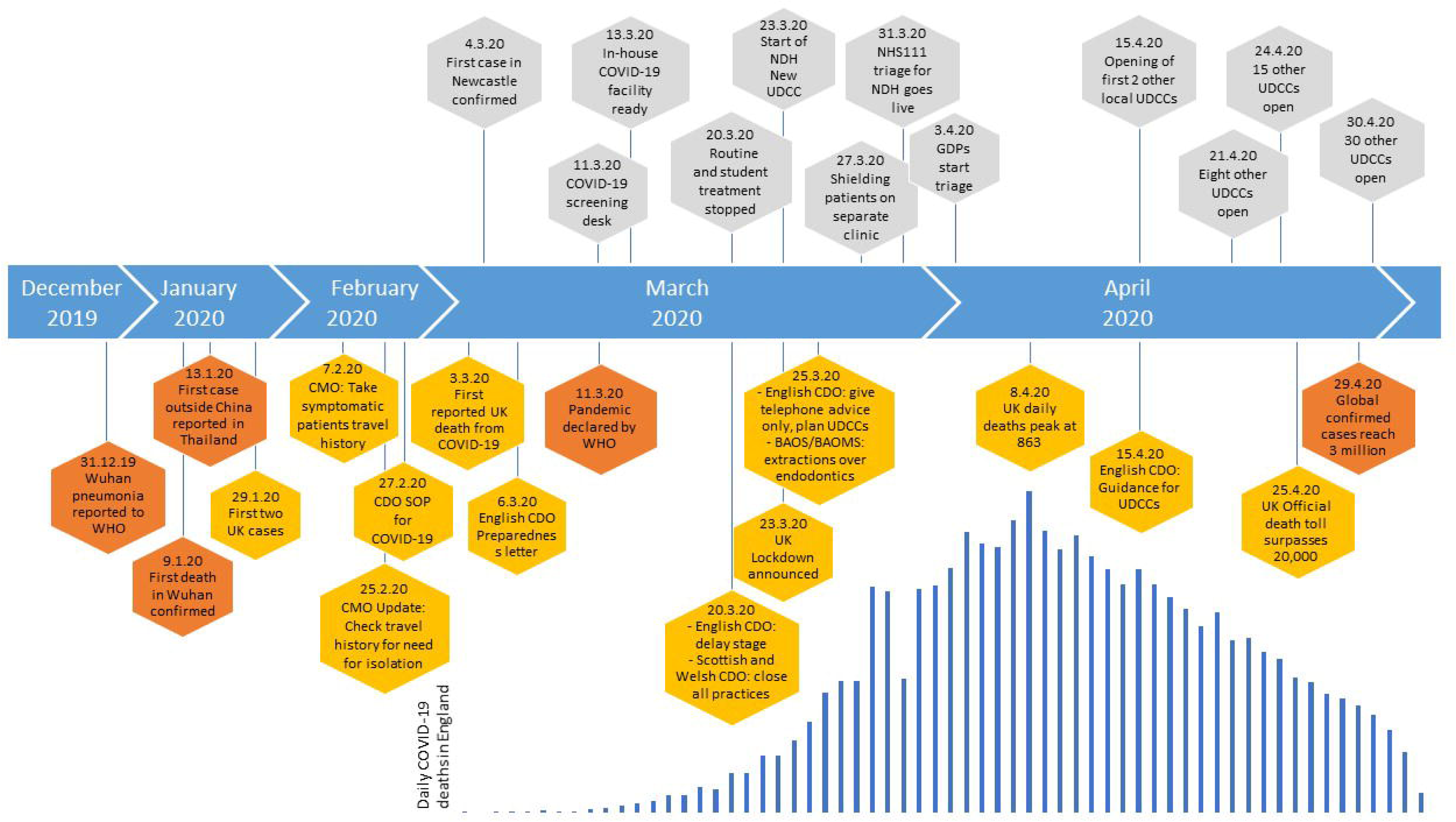
**Description:** Timeline of international (orange), national (yellow) and local (grey) events in the COVID-19 pandemic. UDCC: Urgent Dental Care Centres. CMO: Chief Medical Officer. CDO: Chief Dental Officer. GDPs: General Dental Practitioners. BAOS: British Society of Oral Surgeons. BAOMS: British Association of Oral and Maxillofacial Surgeons. WHO: World Health Organization.

Units from around the world have started to share their experiences of setting up similar UDCCs services in their regions and healthcare settings (10,11). Throughout this pandemic a service evaluation was conducted at NDH UDCC. The aim of the service evaluation was to examine the volume of patient throughput, their demographics and outcomes in order to inform local decision-making and optimise appropriate care pathways. This paper presents our findings from the first 6 weeks and, is one of the first reports in the literature describing and exploring patient characterises and outcomes of those attending an UDCC during the COVID-19 pandemic.

## Methods

A prospective service evaluation was carried out, which was registered on the NUTH Clinical Effectiveness Register (REF 10006) and approved before commencement. Data were collected on all patients attending the NDH UDCC, including paediatric, orthodontic and OOH care. Parameters recorded included: clinic attended, date of attendance, gender, date of birth, partial postcode, referral source, patient GDP registration status, triage type (telephone or face-to-face), triage and clinical consultation diagnoses and outcomes, COVID-19 status, and any repeat attendances at the UDCC. Ethnicity data were also collected retrospectively once it became apparent from media coverage that ethnicity may have an impact on COVID-19 prognosis. Anonymised data were collected by a selection of the UDCC clinicians (RH, AA, RG, GT, CH, SS, CD) using an excel spreadsheet (Microsoft Office Professional Plus 2016, Version: 16.0.4993.1001) from the 23^rd^ March 2020 to 3^th^ May 2020. Aerosol generating procedures (AGPs) were defined as pulp extirpations or surgical extractions (using a handpiece) for the purpose of this service evaluation.

Data cleaning was completed both manually and in STATA release 13 (StataCorp LP, College Station, TX, USA). Where outcomes were identified as being missing clinical notes were reviewed and data added retrospectively where available. Data were analysed in Statistical Package for the Social Sciences (Windows version 25.0.0.1; SPSS Inc., Chicago [IL], US) using descriptive statistics and exploratory analysis with chi-squared tests. Geolytics (https://geo.sg/, accessed 04/05/20) was used to produce geographical location figures. Estimated distance travelled was calculated using the fastest road route as determined by ZIPCodeSof (http://www.postcode-distance.com/distance-between-postcodes). Postcodes more than 210 kilometres away were excluded from the distance travelled analysis on the assumption that these people were not living in their primary residence during the pandemic.

## Results

There were a total of 3068 patient consultations at the UDCC over the 6-week period (1595 telephone, 151 face-to-face and 1322 clinical consultations). The majority of these consultations were for the in-hours adult service (73%, n=2250), with 11% (n=331) being for the in-hours paediatric service and 3% (n=102) for orthodontic services. Thirteen percent (n=385; 90% [n=347] adult, 10% [n=38] paediatric) of consultations were in the OOH service (OOH data includes face-to-face consultations only as triage was provided by an external provider, NHS111/ DCAS).

The distribution of the triage and clinical consultations over the 6-week period is shown in Figure 2. The proportion of triage telephone consultations that resulted in a clinical visit increased during the first three weeks, from 26% (n=37) in week 1 to 51% (n=112) in week 3, before stabilising between 44-51% during weeks 4-6. Face-to-face triage consultations (at the hospital reception desk) were most frequent in weeks one (n=87) and two (n=24) before reducing to very low levels in subsequent weeks (week 3, n=6; week 4, n=17; week 5, n=13; week 6, n=4). The vast majority (94%, n=142) of these face-to-face triage consultations at the hospital reception desk resulted in a clinical visit.

**Figure 2.**
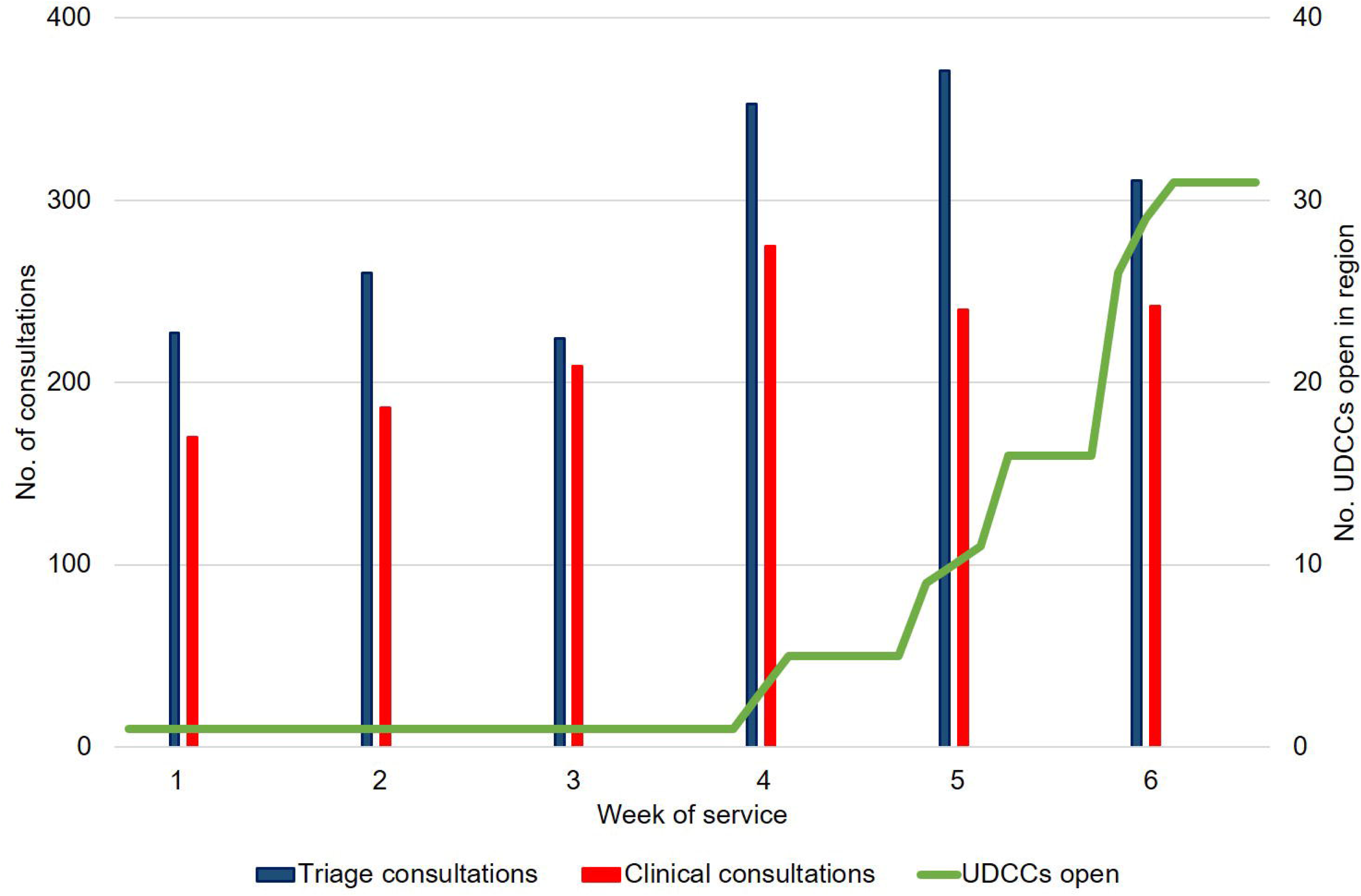
**Description:** Number of triage and clinical consultations by the Newcastle Dental Hospital Urgent Dental Care Centre (NDH UDCC) by week of service during the COVID-19 pandemic. Number of UDCCs open in the region shown for reference (green line). Out-of-hours triage consultations (provided by NHS111) are not included in this figure as the data is unavailable. Week 1 starts on 23^rd^ March 2020, week 6 ends on 3^rd^ May 2020.

The number of clinical consultations increased from week 1 (n= 170) to week 4 (n=275) before stabilising in weeks 5 (n=240) and 6 (n=242). Both genders were equally represented in those accessing the service (50% male and female). The mean age of those accessing the service was 36.5 years +/-19.1 (SD). In terms of patient ethnicity, 76% of patients attending clinical consultations were recorded as White British. Black, Asian and Minority Ethnic (BAME) made up 3.5%, with 19% being other, not known or stated. Detailed patient demographics are summarised in the appendix.

The geographical area served by the NDH UDCC over this period is illustrated in Figure 3. Initially a wide area was served, with patients travelling a mean of 22 km +/-26 (SD; range 0-188 km) to access care during the first week, peaking at 29 km +/-21 (SD, range 3-206) in week 4 and then reducing to 17 km (range 0-79 km) for week 6. The home locations of those patients who accessed telephone triage services and the weekly breakdowns are shown in the appendix.

**Figure 3.**
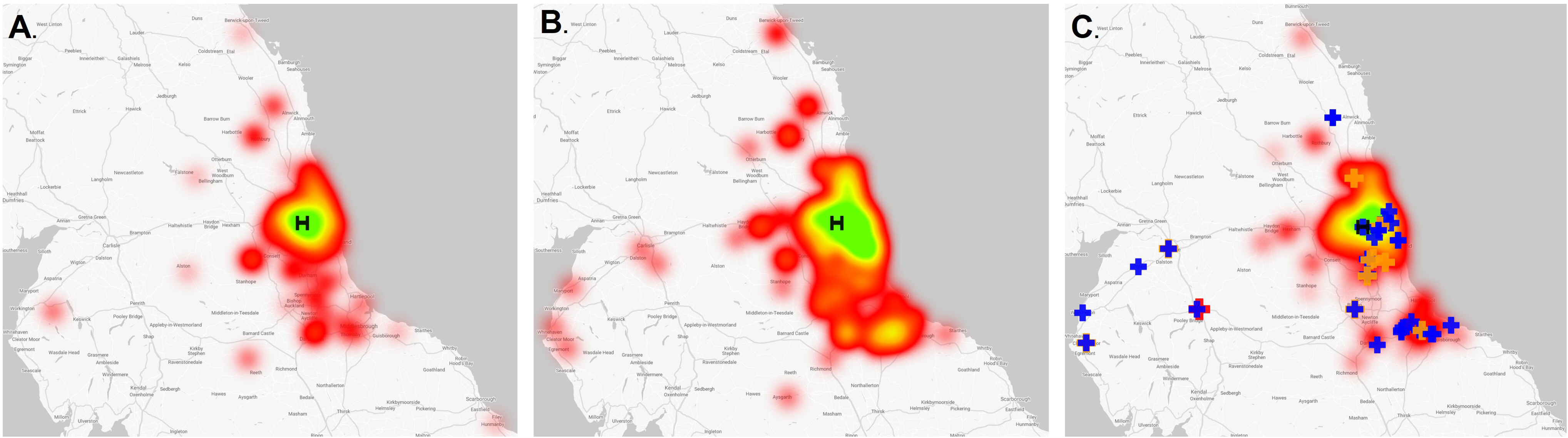
**Description:** Geographical area served by the Newcastle Dental Hospital Urgent Dental Care Centre (NDH UDCC). Heatmap indicates home address of those attending clinical visits in-hours (Out-of-hours data excluded as this service normally covers a wide region). Green = highest signal. Black ‘H’ illustrates the NDH UDCC location. Panel A = weeks 1-2, Panel B = weeks 3-4, Panel C = 5–6. Other UDCC locations shown in panel C by crosses with blue indicating ‘cold’ sites, orange indicating ‘warm’ sites and ‘red’ indicating hot sites.

The majority of patients who accessed the service through the in-hours triage service reported having a GDP (n=1417, 81%) and out of these, 1126 (79%) had attempted to contact them before contacting the UDCC, although this was lowest in the first week (59%) (see appendix). One hundred and thirty (7%) had received no or inadequate advice or triage from their GDP (i.e. provision of AAA, where appropriate), most frequently in the second week (n=78, 30%), however this markedly improved in later weeks (see appendix). Two hundred and twelve (12%) patients reported not having a GDP they saw regularly for treatment prior to the pandemic. There was a significant association between gender and having a GDP with males significantly more likely to report that they did not have a GDP (X^2^ (1, *n* = 1628) = 8.204, *P*=0.004).

The diagnoses of triage and clinical consultations is shown in figure 4 (additional data including weekly breakdowns is provided in the appendix). Most attendances were for acute pulpal and periapical complaints, which was consistent across adult, paediatric and OOH services. Symptomatic irreversible pulpitis was the most common triage diagnosis, whereas symptomatic apical periodontitis was the most common clinical diagnosis. The outcome of the clinical consultations is shown in figure 5, with the majority resulting in extractions (63%, n=832); only 0.5% (n=8) were AGPs. Sixty three percent of extracted teeth were molars, 15% premolars, 5% incisors/canines, with the remaining being primary teeth or multiple paediatric extractions.

**Figure 4.**
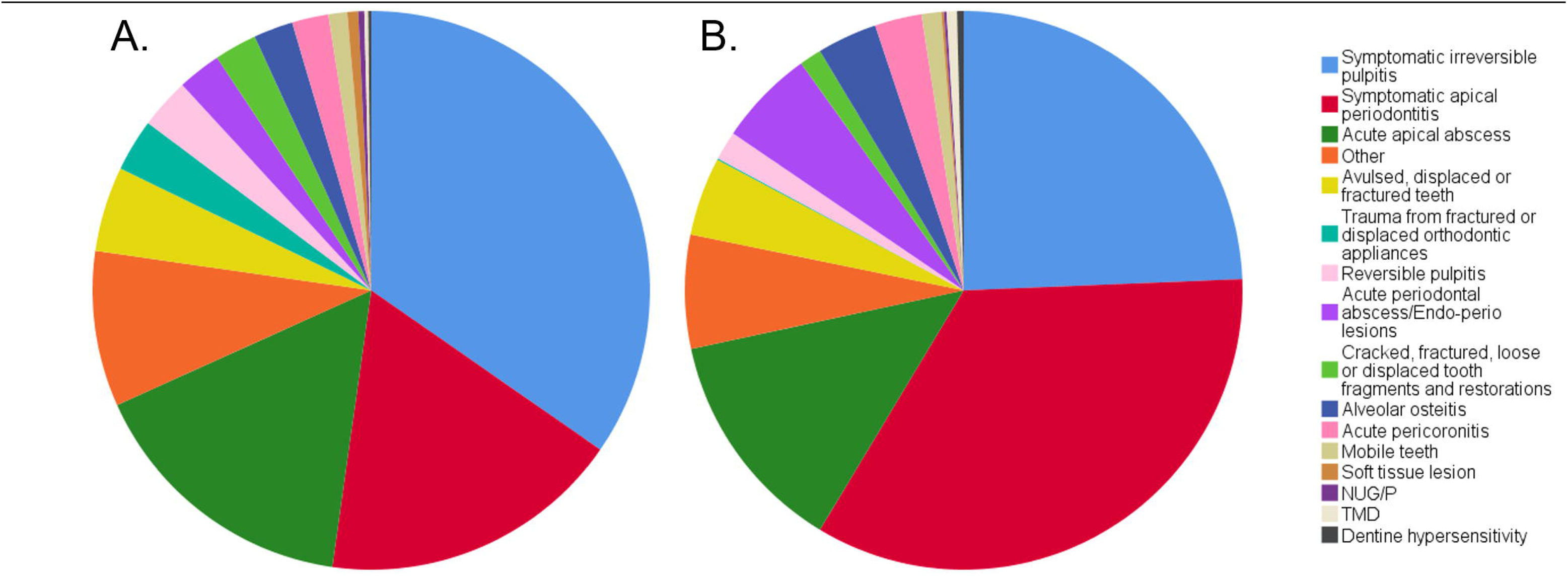
**Description:** Diagnoses of triage (A) and clinical (B) consultations in the Newcastle Dental Hospital Urgent Dental Care Centre (NDH UDCC) by week of service during the COVID-19 pandemic. Out-of-hours triage consultations (provided by NHS111) are not included in this figure as the data is unavailable. ‘Other’ notable diagnoses included: post-extraction pain (triage, n=29; clinical, n=11), removable prosthesis problems (triage, n=3; clinical, n=1) and extra-oral non-odontogenic swelling (triage, n=2; clinical, n=1).

**Figure 5.**
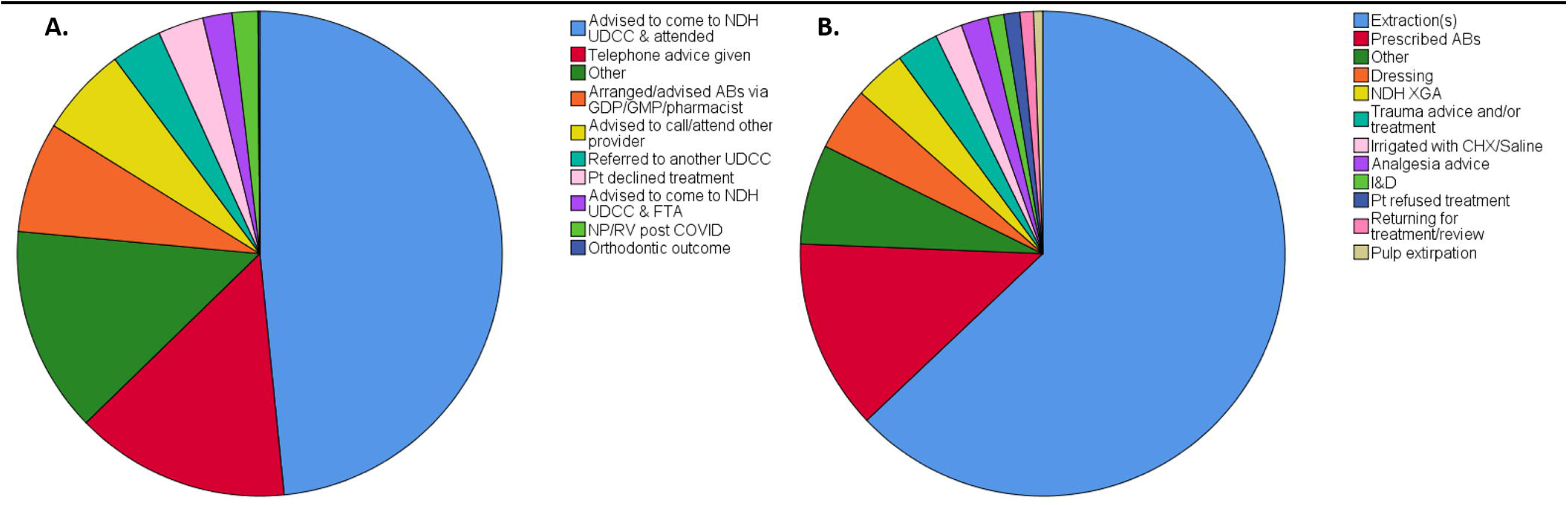
**Description:** Outcomes of triage (A) and clinical (B) consultations in the Newcastle Dental Hospital Urgent Dental Care Centre (NDH UDCC) by week of service during the COVID-19 pandemic. For triage outcomes, those who were asked to call/attend other providers this 89% (n=93) to GDPs, 9% (n=9) to NHS111 and 2% (n=2) to A&E. Telephone advice constituted 85% (n=213) receiving analgesia advice, 8% (n=19) receiving trauma advice and 7% (n=18) advised temporary filling kit. For clinical outcomes, 3% (n=22) of the extraction outcomes had antimicrobials also prescribed, whilst 2% (n=17) were incomplete extractions i.e. root or apex fracture with follow up arranged.

Of the patients attending clinical consultations, and with a triage diagnosis indicating AAA was suitable, over half (n=245, 56%) had completed AAA, 35% (n=153) had only received advice and analgesia advice but no antimicrobials and 5% (n=21) had received no advice. There was a gradual increase in the number of patients attending the clinical consultations with failed AAA, when the triage diagnosis suggested this was a suitable strategy, from 30% in week 1 to 68% in week 4 (full details in appendix). For those with a triage diagnosis of symptomatic irreversible pulpitis and attending a clinical visit, 48% (n=183) had received AAA, 43% (n=164) AA and 6% (n=21) no advice.

The vast majority of patients attending the service were asymptomatic and non-shielding for COVID-19 (79%, n=1043), 15% (n=200) were shielding, and 1% (n=17) were symptomatic. The proportion of shielding patients increased, from a level of 2% (n=4) in week 1, to a peak of 21% (n=57) in week 4. Eight (0.5%) of those triaged via the telephone as having a COVID-19 status of asymptomatic or shielding were subsequently deemed symptomatic and moved to the appropriate department or asked to go home and return after their isolation period. The weekly breakdowns and COVID-19 status of the triage consultations is shown in the appendix.

The majority of patients attending clinical consultations were first-time attenders (86%, n=1137), although 7% (n=96) had previously attended during the COVID-19 pandemic and 5% (n=60) prior to the pandemic. Of those returning for repeat visits during the pandemic the majority were for alveolar osteitis (42%, n=40) or for continuing acute pulpitic or periapical symptoms/diagnoses. The rate of patients returning with alveolar osteitis (40 cases / no. extraction-based procedures in the first 5 weeks) was 6%.

## Discussion

This service evaluation played an important role in the evolution and quality improvement of the NDH UDCC. It informed triage processes, staffing levels and provided useful feedback to our partners at the NHS111 service, DCAS and GDPs. The data allowed the UDCC to use service information to inform decisions, alongside the guidance being produced. The data also gives a unique opportunity to record and document urgent dental care demands when primary dental care services close, leaving only one provider, as was the case for the first three weeks of this service. The insights from this data will help inform future pandemic preparedness planning.

Over the 6-week period there was a relatively equal split of patient gender attending the UDCC, this is in contrast with typical patient demographics accessing emergency dental care whereby the patient population is predominantly male (12–14). This change in demographic could reflect the change in service provision, whereby female patients who were more likely to be registered with a GDP in this cohort, and who are more likely to seek care at an early stage (15,16) started to present to the UDCC rather than primary care. The mean patient age was 37 years, which is in keeping with the literature of emergency dental care attenders who are typically in the third or fourth decade (12–14,17,18). The majority of patients attended with diagnoses of symptomatic irreversible pulpitis or apical periodontitis, which is again in keeping with the literature on emergency dental care (12,17,19–21).

Over half of the patients seen for clinical consultation had followed national guidelines with regards to GDP triage and AAA provision, where this was appropriate, prior to attending. Use of antibiotics for dental pain during the pandemic is a controversial area (22), as on one hand UDCC attendances needed to be minimised, whilst also remaining aware of the potential for antibiotic resistance to increase with inappropriate use, the continued paucity of evidence to support their use in the management of acute localised dental painful conditions (23) and the potential for adverse side effects. The number of patients reporting failure of AAA increased throughout the study period. This increase could represent more stringent enforcement of optimal AAA by our triage team prior to a clinical visit, or it could suggest that these measures aren’t sufficient to manage severe dental pain. Advising use of antibiotics in the management of any acute dental problem remotely without the benefit of a clinical examination is challenging for clinical staff. Given the absence of clinical examination it could risk inappropriate prescribing and thereby delivers a confusing public health message following several years of campaigning that “antibiotics do not cure toothache”(24). However, given the requirement to immediately stop dental care, and without arrangements in place for urgent care provision, the AAA strategy does seem to have been the only option in this initial stage, but could, with the benefit of hindsight, have been colloquialised as the 4A approach – Advice, analgesia and if appropriate antibiotics thereby emphasising that antibiotics would only be appropriate in certain circumstances. Future preparedness planning should take this into account and hopefully avoid the need for a repeat of this situation.

Demand for the NDH UDCC service was high, with an initial peak of telephone calls immediately following the publication of the Chief Dental Officer for England’s letter directing all primary care GDPs to give telephone advice only (3). This was followed by a continued high number of referrals to the service, which peaked in week 5 before declining as more UDCCs became operational across the region in weeks 5 and 6. Due to the initial almost four week period where NDH UDCC was the only service provider, patients accessed the service from a wide geographic area. This was clearly an undesirable situation given that UK government advice recommended only essential travel, avoidance of public transport where possible (25), and the need for community-centred care to reduce flow into hospitals in order to limit the spread of infection (26). In the final two weeks as more UDCCs were established, patient volumes decreased and patients became more local to the hospital, allowing a reduction in staffing level on the clinic and in turn, the number of potential infectious contacts for staff members. This demand for the NDH UDCC in the early stages of the pandemic, due to lack of service provision regionally, demonstrates the need for early contingency planning in dentistry for future pandemics. Our data may inform the management of future changes to routine dental care during this pandemic, or the next.

Fortunately, the majority of patients attending the UDCC centre were asymptomatic for COVID-19. This may be because symptomatic patients were following UK government guidelines and were self-isolating, too unwell with COVID-19 to consider dental pain a priority, or may also indicate the relatively low numbers of patients who coincidently had COVID-19 infection and require UDCC treatment. Our management of symptomatic or isolating patients was to try and avoid any attendance if at all possible, accepting only dental trauma, swelling or bleeding as reasons for attendance in this group, which would have reduced this number further. A challenging diagnostic dilemma was in the management of pyrexic patients attending with dental abscesses. In absence of further information, a cautious approach was utilised in this circumstance as we had sufficient capacity and resource to do so.

The NDH UDCC was staffed by 69 clinicians and 55 dental nurses on a rotational basis. The in-hours adult UDC service comprised 38 clinicians and 30 dental nurses. To the best of our knowledge no staff acquired infection occurred due to occupational exposure. We adopted the BAOS/BAOMS guidance advocating sessional use of FFP3 masks (6). Although this is beyond the level of PPE recommended by PHE, NDH felt that, initially, as the only provider of dentistry in the region, with staffing and patient footfall to match this, there was an increased risk of staff becoming infected and increasing the risk of nosocomial infection for patients and other staff (26). NUTH had good capacity for staff testing early on during the pandemic, and have found that staff in patient-facing roles had a similar infection rate to those who work in administrative/back of house areas, suggesting community acquired infection with COVID-19, rather than occupational (27), supporting the current management of PPE within the trust.

The most common treatment was extraction, with the majority of teeth being posterior teeth. This was in line with our local agreement to avoid AGPs unless essential, e.g. to restore strategically important teeth only. The presence of open dental pulps and oral sepsis is known to be more prevalent in lower socioeconomic groups (28), and the area we serve includes some of the most deprived communities in England (29). This may account for a high extraction rate as a result of the extent of disease at presentation to the UDCC alongside our planned strategy of carrying out AGPs only when deemed necessary.

There are limitations of our data which were collected as part of an ongoing service evaluation and hence in a pragmatic approach. There was no formal calibration of the data collectors, however they were all experienced clinicians and were able to communicate easily within the team for any queries and consensus regarding data collection and input.

The COVID-19 status data should be interpreted in the context of the testing limitations in the UK during this period (limited to symptomatic patients admitted to hospital, or symptomatic key [health and social care] workers) and hence this status relied on the presence or recent history of symptoms. Data were collected from existing documentation and on occasions this has meant there are missing data in some cases. For the ‘triage diagnosis’ this was not always recorded and in this case the data collectors were asked to review the information recorded and use their judgement to give the most likely diagnosis (or reason for contact). Our anonymised data collection did not include a patient identification number meaning we were unable to retrospectively state the exact number of individual patients seen within the service, rather we report consultation episodes. The data and experiences presented in this paper are our experience. They will be influenced by geographical, cultural and organisational factors which needs to be considered when generalising them to other locations and settings.

## Conclusion

In conclusion, our data gives an insight into the patient characteristics, including urgent dental care requirements, of a lone regional provider during the COVID-19 pandemic crisis. A telephone triage service was essential, and demand grew over time. Acute pulpitic or periapical symptoms were most common and most AGPs could be avoided. Very few symptomatic COVID-19 patients accessed the service but demand from those shielding was high. Dental preparedness for this pandemic crisis at national, regional and local levels, was challenged and these data will help inform future planning.

## Data Availability

Data available in article supplementary material or by request from the authors

## Acknowledgements

R.Holliday and C.Currie are funded by the National Institute for Health Research. This paper presents independent research funded by the National Institute for Health Research (NIHR). The views expressed are those of the authors and not necessarily those of the NHS, the NIHR or the Department of Health. The authors declare no potential conflicts of interest with respect to the authorship and/or publication of this article.

Thanks to all the staff at NDH for their support in managing the extensive service changes in this time, in particular but not exclusively, Lee Mercer, Kelly Gillan, Jenna Trainor, Phillipa Graham, Marie Allen, Jan McCalister, Andy Pike, Stewart Youngman, Tom Robson. We would also like to thank our local primary dental care colleagues who have been flexible and understanding during this challenging period.

## Author Contributions

E. Carter, C.Currie, R.Holliday, contributed to the conception, design, data acquisition, and interpretation, drafted and critically revised the manuscript; J.Durham, G.Walton, B.Cole, M.Greenwood, contributed to the data interpretation, drafted and critically revised the manuscript; A.Asuni, R.Goldsmith, G.Toon, C.Horridge, S.Simpson, C.Donnell, contributed to the data acquisition, drafted and critically revised the manuscript.

